# Analysis of Epidemic Situation of New Coronavirus Infection at Home and Abroad Based on Rescaled Range (R / S) Method

**DOI:** 10.1101/2020.03.15.20036756

**Authors:** Xiaofeng Ji, Zhou Tang, Kejian Wang, Xianbin Li, Houqiang Li

## Abstract

1

**Background:** The outbreak of the new coronavirus infection broke out in Wuhan City, Hubei Province in December 2019, and has spread to 97 countries and regions around the world. Apart from China, there are currently three other severely affected areas, namely Italy, South Korea, and Iran. This poses a huge threat to China’s and even global public health security, challenges scientific research work such as disease surveillance and tracking, clinical treatment, and vaccine development, and it also brings huge uncertainty to the global economy. As of March 11, 2020, the epidemic situation in China is nearing its end, but the epidemic situation abroad is in the outbreak period. Italy has even taken measures to close the city nationwide, with a total of 118,020 cases of infection worldwide.

**Method:** This article selects the data of newly confirmed cases of COVID-19 at home and abroad as the data sample. Among them: the data of newly confirmed cases abroad is represented by Italy, and the span is from February 13 to March 10. The data of newly confirmed cases at home are divided into two parts: Hubei Province and other provinces except Hubei Province, spanning from January 23 to March 3, and with February 12 as the cutting point, it”s divided into two periods, the growth period and the recession period. The rescaled range (R / S) analysis method and the dimensionless fractal Hurst exponent are used to measure the correlation of time series to determine whether the time series conforms to the fractal Brownian motion, that is, a biased random process. Contrast analysis of the meaning of H value in different stages and different overall H values in the same stage.

**Results:** Based on R / S analysis and calculated Hurst value of newly confirmed cases in Hubei and non-Hubei provinces, it was found that the H value of Hubei Province in the first stage was 0.574, which is greater than 0.5, indicating that the future time series has a positive correlation and Fractal characteristics; The H value in the second stage is 1.368, which is greater than 1, which indicates that the future epidemic situation is completely preventable and controllable, and the second stage has a downward trend characteristic, which indicates that there is a high probability that the future time series will decline. The H values of the first and second stages of non-Hubei Province are 0.223 and 0.387, respectively, which are both less than 0.5, indicating that the time series of confirmed cases in the future is likely to return to historical points, and the H value in the second stage is greater than that in the first stage, indicating that the time series of confirmed cases in the second stage is more long-term memory than the time series of confirmed cases in the first stage. The daily absolute number of newly confirmed cases in Italy was converted to the daily growth rate of confirmed cases to eliminate the volatility of the data. The H value was 1.853, which was greater than 1, indicating that the time series of future confirmed cases is similar to the trend of historical changes. The daily rate of change in cases will continue to rise.

**Conclusion:** According to the different interpretation of the H value obtained by the R / S analysis method, hierarchical isolation measures are adopted accordingly. When the H value is greater than 0.5, it indicates that the development of the epidemic situation in the area has more long-term memory, that is, when the number of confirmed cases in the past increases rapidly, the probability of the time series of confirmed cases in the future will continue the historical trend. Therefore, it is necessary to formulate strict anti-epidemic measures in accordance with the actual conditions of various countries, to detect, isolate, and treat early to reduce the base of infectious agents.

## 2 Introduction

At the beginning of the outbreak of the new coronavirus infection, some cases were related to the South China Seafood Market in Wuhan City, Hubei Province. At first, it was diagnosed as “unknown cause of pneumonia”, but it quickly spread to all parts of the country and parts of Southeast Asia, North America, Europe and other countries and regions. It is a virus that can develop into severe respiratory diseases. On February 11, 2020, the International Virus Classification Commission officially named the virus “Severe Acute Respiratory Syndrome Virus 2”, referred to as “SARS-CoV-2”. At the same time, the World Health Organization named the virus-infected pneumonia “Coronavirus 2019 (COVID-19) “and escalated the epidemic into an international public health emergency of concern. As of March 11, 2020, a total of 118,020 COVID-19 diagnoses have been confirmed worldwide. A total of 37,064 cases have been confirmed abroad, of which 7,755 cases have been confirmed in South Korea and 10,149 cases have been confirmed in Italy. A total of 80,956 cases have been confirmed in China, of which 67,73 cases have been diagnosed in Hubei Province. The origin of the new coronavirus has not yet been determined, and various evidences suggest that its source of infection comes from wild animals. The medical field mainly focuses on three aspects: etiological characteristics, clinical diagnosis, and epidemiology, Several studies have shown that SARS-CoV-2 is most similar to bat SARS-like coronaviruses (SARS-like CoVs) and is more infectious than SARS viruses during the incubation period.^[1]^

Since the outbreak of the new crown virus infection in Wuhan in 2019, many scholars have performed simulation analysis on the spread and prevention of the epidemic, and reached different conclusions. Ziff (2020) believes that using the basic infection number R_0_ to measure the possibility of large-scale transmission of the virus without prevention and control is not realistic.^[2]^ The main drawback is that it is necessary to assume that the virus”s propagation path is exponential, and it can be corrected using the SEIR logarithmic model.^[3]^ Li et al. (2020) used Log-normal, Weibull, and Gamma distributions to estimate the basic regeneration number R0 as 2.2 (95% CI, 1.4 to 3.9) and the latency period to be 5.2 days (95% CI, 4.1-7). The number of confirmed cases doubles every 7.4 days, and the interval from onset to inpatient treatment and cure was 9.1 days (95% CI, 8.6-9.7). ^[4]^ Wu et al. (2020) analyzed and predicted by SEIR model and estimated the basic regeneration number R0 to be 2.6 (95% CI, 2.47-2.86), the estimated cumulative total confirmed cases was 75,815 (95% CI, 37304 to 130330) and the number of confirmed casesDoubled at 6.4 days (95% CI, 5.8-7.1). ^[5]^ The calculation shows that the transmission path of New Coronavirus has certain fractal process characteristics. By comparing the difference between the actual confirmed case and the theoretical value, you can determine which period the epidemic is in:rising or falling. Vazquez (2006) uses the branching process of fractal theory to describe the spread of variables, to establish mathematical models for different situations such as population or virus transmission in reality, to determine the corresponding power index and to calculate the potential development path of historical data.^[6]^ Throughout the severe situation of the global economy, the spread of the new crown virus infection epidemic has led to a sudden drop in demand and production, which has affected the global supply chain economy. In the external environment of economic downturn, how to maintain macroeconomic stability and find opportunities among risks has become the focus of attention. ^[7]^

## 3 Rescaled Range (R / S) Analysis

### 3.1 Overview of R/S and H value theory

The R/S analysis method was discovered by the British hydrologist H.E.Hurst in 1951 when he examined the Nile discharge variation. ^[8]^ It is a widely used robust non-parametric statistical method. The biggest advantage is that the measured time series does not need to assume its specific distribution characteristics whether the sequence is smooth. The robustness of the R/S analysis results is not affected by the distribution characteristics.^[9]^

H value is also called Hurst index. Hurst believes that the following relationship exists between R / S and H: (R / S) t = (t / 2) H, which can be obtained by log (R / S) and log (n) regression fitting for judgment Trend and randomness of time series Whether the time series is a fractal B-rownian motion is determined according to whether the H value is equal to 0.5. The H index has a wide range of applications. Cajueiro (2004) believes that the H index can be used to determine the effectiveness of the securities market. This method can be extended to other fields that study the d-ynamic characteristics of time series. ^[10]^ When predicting a set of memorable data, it can refer to its historical development path. Improper selection of the length of the time series will affect the bias of the estimation of the H index and reduce the accuracy of the prediction. To correct this error, Barunik (2010) uses a rolling time window to deter-mine the form of the H index over time, and uses the AR (1) -GARCH (1,1) process to filter the s-equence. Multifractal detrending analysis (MF-DFA) uses different polynomials to fit different ord-er moments of variables. It is a generalization of the DFA method and can improve the accuracy o-f processing thick-tailed data.^[11]^

### 3.2 Model building

In this paper, the rescaling range method is used for modeling. It is assumed that the transmission process of the New Coronavirus conforms to the expansion path of fractal theory. At the same time, Italy uses the newly diagnosed change rate. The R/S value is linearly regressed on the time series span to obtain the H index of different regions, determine the fractal dimension of the model, and analyze the possible change direction of historical data over time.

Hypothetical time series{x _i_}, *i* = 1,2,3⋀, *n* the span is the time series length n.Segment this time series to construct n-t+1 subsequences. Let the divided subsequence be {*Y* _*j*_ }, *j* = 1,2,3⋀, *n* − *t* + 1,Where t is the length of the initial segmentation subsequence, and the last subsequence is the time series itself. The relevant information of each subsequence is shown in Table 1.

**Table1.** Subsequence for time series segmentation

| Subsequence number | Subsequence length | The data contained in the subsequence |
| --- | --- | --- |
| 1 | t | $X_1, X_2, \dots, X_t$ |
| 2 | t+1 | $X_1, X_2, \dots, X_t, X_{t+1}$ |
| ..... | ..... | ..... |
| n-t+1 | n | $X_1, X_2, \dots, X_n$ |

Define the mean of the time series as :

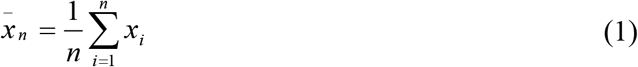

Cumulative dispersion as :

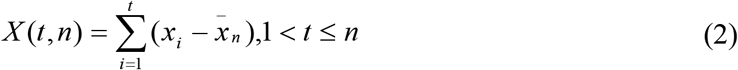

The range as :

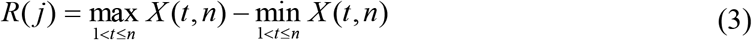

The mean square error of the time series as :

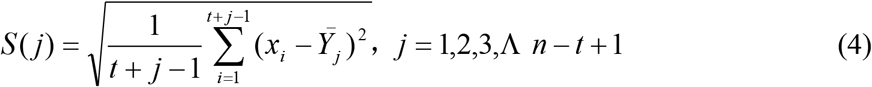

Among them 

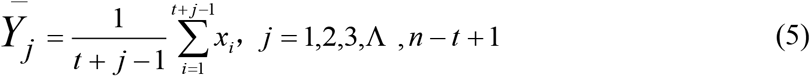

Rescaled R / S as: 

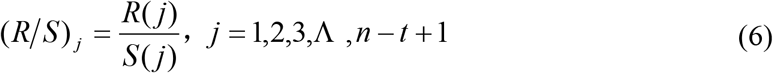

With respect to each subsequence of length, n-t+1 range and mean square errors can be obtained. The Hurst exponent can be approximated by plotting the image with log(R/S)_j_ and log(t). where the base of the logarithm is 10, and the approximate value of Hurst is the slope of the linear regression model fitting curve. 

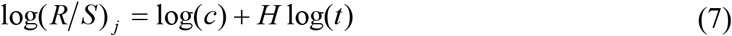

Where c is a constant. The Hurst index of the time series can describe the trend characteristics of the self-similarity of the time series. When 0 <H <0.5, the time series has anti-persistence, that is, the future change increment of the time series is opposite to the change increment of the historical period; when H = 0.5, the standard Brownian motion, the time series is random walk; when H> 0.5, the future data growth trend of the time series is similar to a certain historical trend, with positive persistence, long-term memory and fractal characteristics.^[12]^

## 4 Predictability analysis of new coronavirus infections

### 4.1 The data description

This article analyzes the epidemic situation in China in two stages: the first stage is from January 23 to February 11; the second stage is from February 13 to March 3. The subjects selected are also divided into two: one is the newly confirmed daily cases in Hubei Province; the other is the total daily newly confirmed cases in all provinces except Hubei Province. The newly confirmed case in Italy are highly similar to those in Hubei Province in the early stage, so the main body selected for the analysis of the foreign epidemic was Italy”s newly confirmed cases from February 22 to March 7. The data comes from China”s National Health Committee and the International Health Organization.

### 4.2 Predictability analysis

This article assumes that the length of the initial subsequence of the time series formed by Hubei Province and other provinces except Hubei Province is t=5, and the initial subsequence of the time series formed by the daily rate of newly added diagnostic changes in Italy is t=4. The model analysis is mainly divided into three parts: R/S analysis and calculation of Hurst value for daily new cases in Italy, Hubei Province and other provinces in China. Quantitative methods such as traditional dynamics models, Logistic functions, and Log-normal distributions, due to the different logic of the estimated parameters, result in predictions that differ greatly from actual data. The R/S analysis method does not need to assume the distribution characteristics of the series, and makes a theoretical basis for the future trend change of the time series by estimating the H value.

#### 4.2.1 Analysis of epidemic situation abroad

Because the daily number of newly diagnosed patients in Italy varies greatly, in order to reduce the volatility of the time series, the daily rate of new cases is selected as the time series. Analyze and fit according to the data published daily by the World Health Organization and the China Health Committee, as shown in Figures 1 to 3:

**Figure 1.**
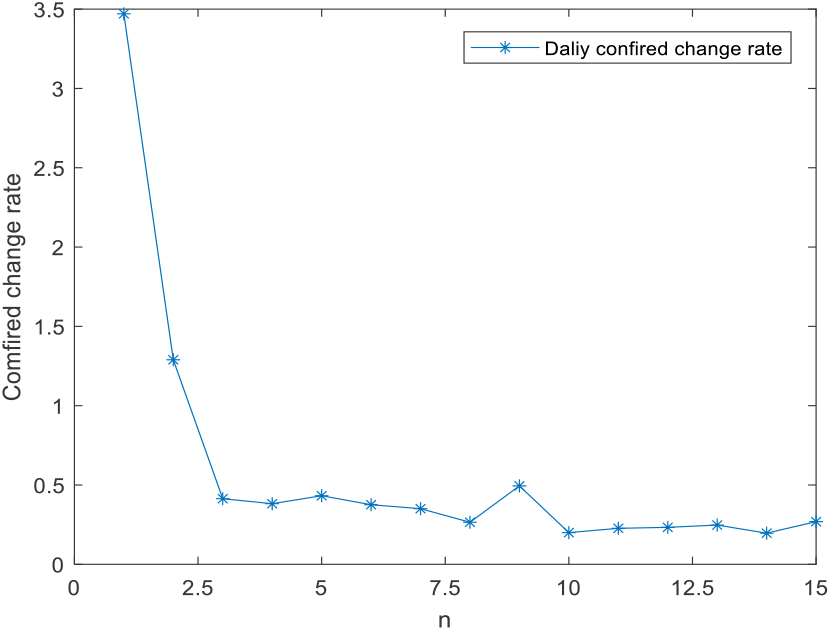
Time series of newly added diagnosed changes in Italy

**Figure 2.**
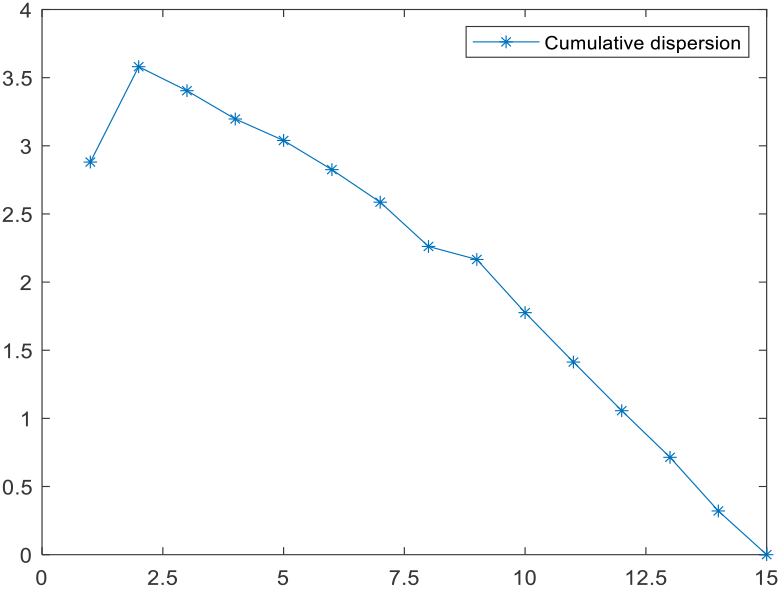
Cumulative deviation (Italy)

**Figure 3.**
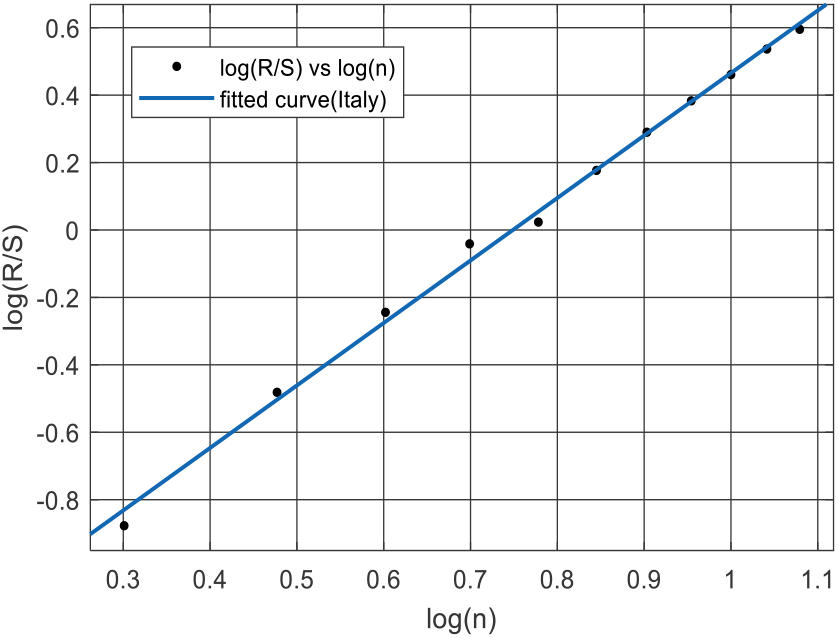
Log(R/S) and log(n) regression fitting (Italy)

##### 1. Cumulative dispersion

The cumulative dispersion reflects the change of the time series data of confirmed cases relative to the time series mean. Observing the cumulative dispersion curve (Figure 2) and the daily increase rate curve (Figure 1), it can be known that when the cumulative dispersion curve is tilted up ward, the data at the corresponding position in the time series is greater than the average value of the time series; When tilting down, the data at the corresponding position in the time series should be less than the average of the time series. In the cumulative dispersion curve (Figure 2), the cumulative dispersion curve formed by the 1-2th data point slopes upward, indicating that the 1-2th data value in the time series (Figure 1) is greater than the average; from Figure 1 and Figure It can be seen that the 3rd to 13th data values are smaller than the average value of the time series.

##### 2. Fractal Hurst Index

Using Matlab to linearly fit log(R/S) and log(n), the regression equation log (R/S)=1.853log (n)-1.388 is obtained. The regression equation shows that the H value is 1.853, which indicates that the time series of the daily rate of newly diagnosed changes in Italy is not a standard Brownian motion and is completely predictive, indicating that the daily rate of newly diagnosed changes in Italy in the short term is similar to the historical trend. The rate of new diagnoses will continue to rise.

#### 4.2.2 Analysis of domestic epidemic situation

4.2.2-1 Analysis of epidemic situation in Hubei Province

The first case of new coronary pneumonia occurred in Wuhan City, Hubei Province.

Analyzing the newly diagnosed patients in Hubei Province and calculating the H value is of reference and predictive significance to countries and regions that are in the outbreak period. This article counts the daily number of newly diagnosed new types of coronary pneumonia in Wuhan from January 20th to March 3rd. Among them, Hubei Provincial Health and Construction Commission changed the authentication method of confirmed cases, which resulted in the sudden increase of new coronary pneumonia to 14840 on February 12th. Data for that day. By observing the characteristics of the time series, the sequence was divided into two sections with February 12 as the dividing point, January 20-February 11 as the epidemic rise phase, and February 13-March 3 as the epidemic decline phase. Figures 4-9 can be obtained by data analysis and fitting. Analysis shows:

**Figure 4.**
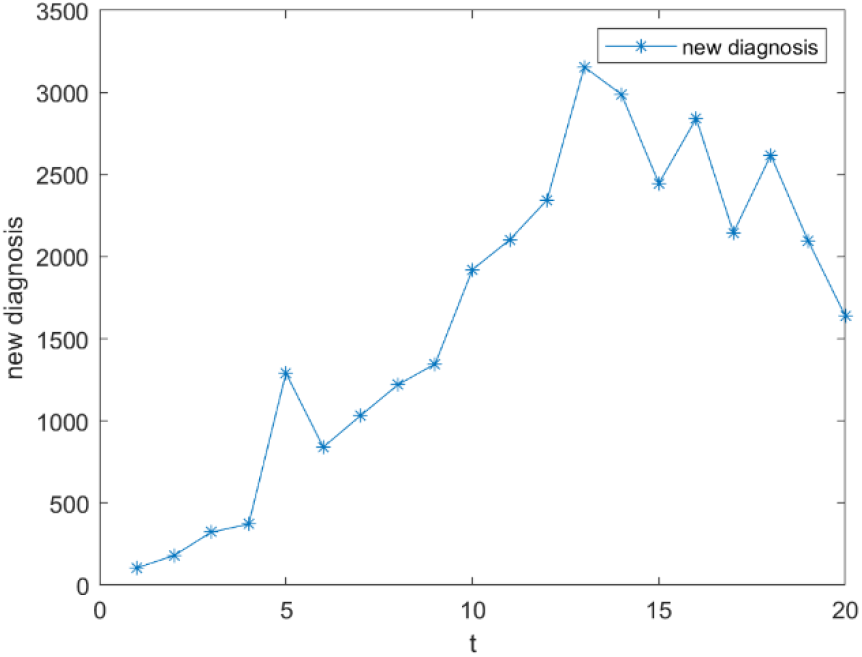
Time series of newly diagnosed patients in Hubei Province (first stage)

**Figure 5.**
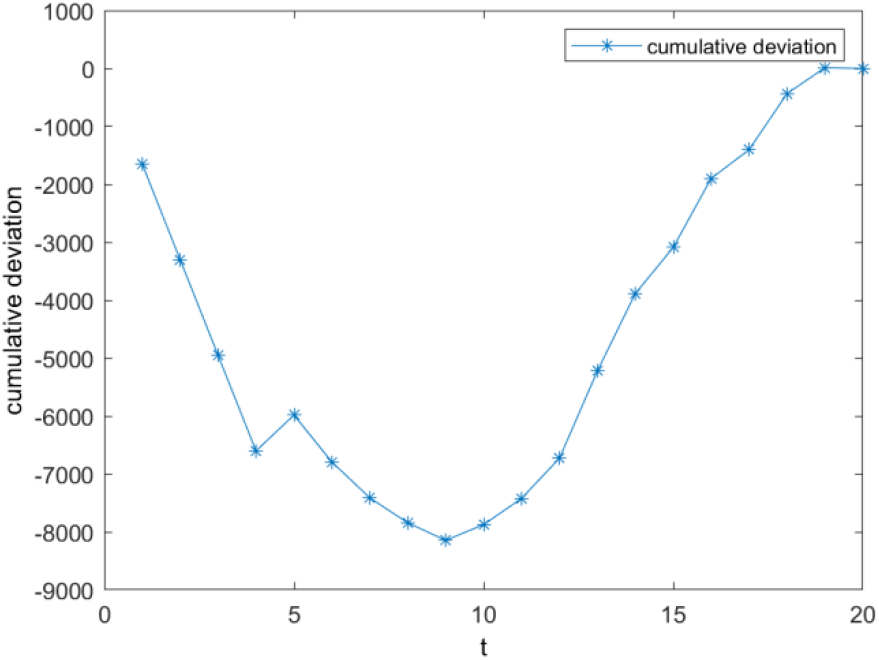
Cumulative dispersion (first stage in Hubei Province)

**Figure 6.**
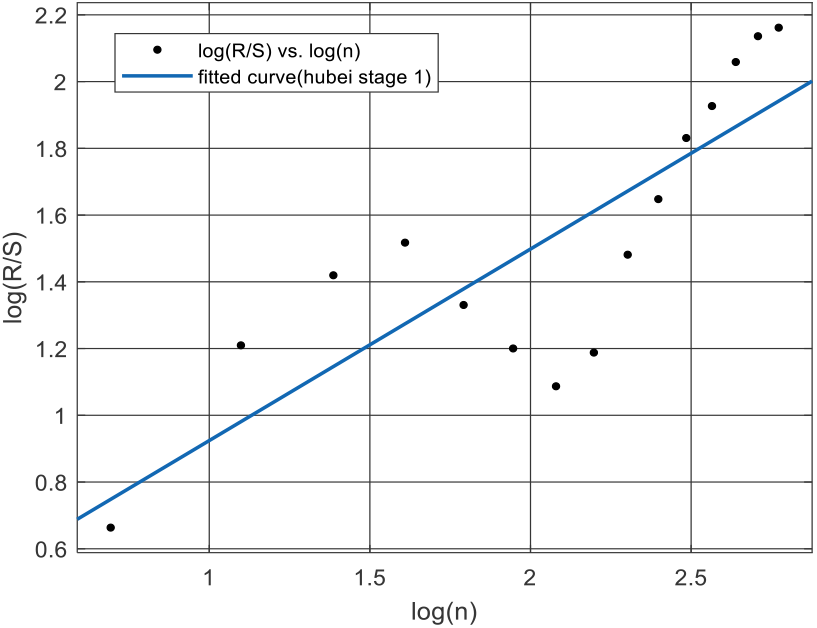
Log(R/S) and log(n) regression fitting (the first stage of Hubei Province)

**Figure 7.**
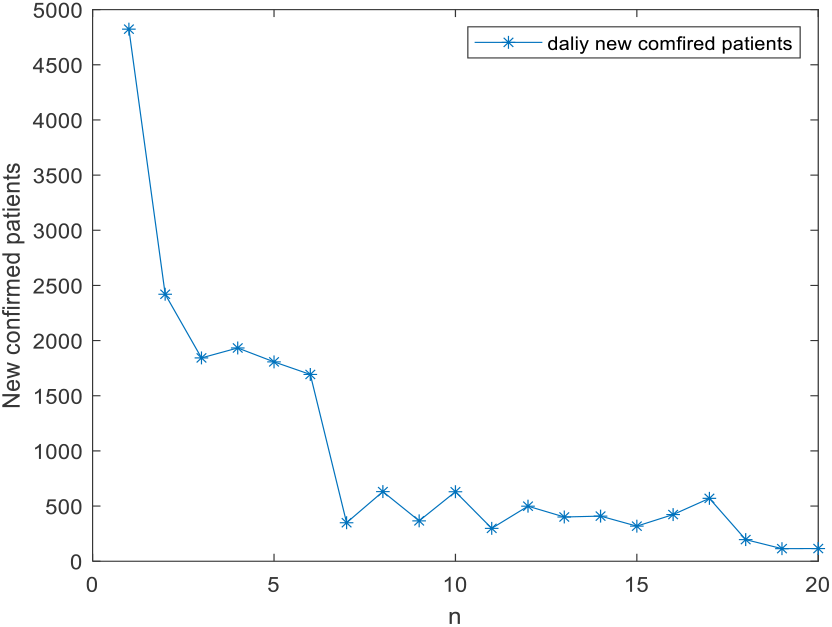
Time Series of Daily New Population in Hubei Province (Second Stage)

**Figure 8.**
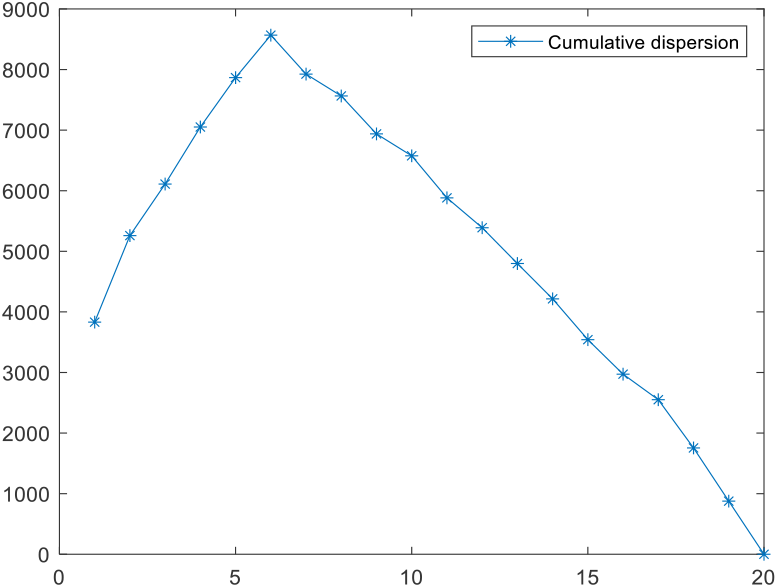
Cumulative dispersion (second stage in Hubei Province)

**Figure 9.**
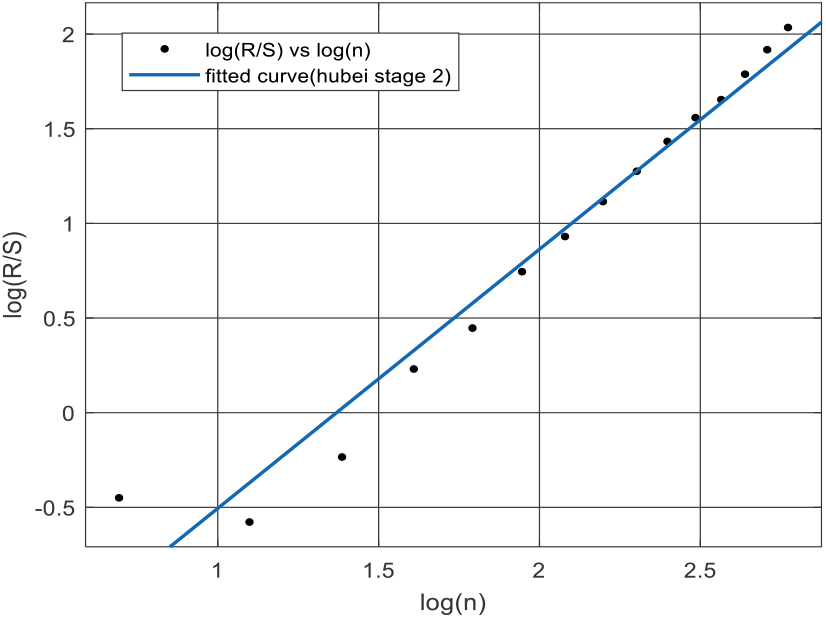
Log(R/S) and log(n) regression fitting (second stage in Hubei Province)

##### 1. Time series characteristics

It can be seen from Figure 4 that the number of newly diagnosed patients on February 4 reached the peak of the first stage, and it seems to have shown a downward trend since then. However, in combination with the number of confirmed patients at the early stage of the second stage of Figure 7 and the data of February 12 that have been removed, we can see that The downward trend in the number of people is an illusion due to imperfect judgment standards. In the second stage (Figure 7), the number of newly diagnosed patients decreased greatly from February 13 to 19, and the number of newly diagnosed patients fluctuated around the value of 400 for more than ten days. Epilogue stage.

##### 2. Cumulative dispersion

The time series curve and cumulative dispersion curve of newly diagnosed patients have associated characteristics. Comparing Figure 4 and Figure 5, it can be seen that in the first stage, the number of newly confirmed patients before January 31 is less than the serial average, and the number of newly confirmed patients from January 31 to February 11 is larger than the serial average. Comparing Figures 7 and 8, it can be seen that in the second stage, the number of newly confirmed diagnoses before February 18 is above the serial mean, and the number of newly confirmed diagnoses from February 19 is below the serial mean.

##### 3. Fractal Hurst Index

Using Matlab to linearly fit log(R/S) and log(n), we can get the first-stage regression equation of Hubei Province log(R/S)=0.574log(n)+0.152. The fitted curve is shown in Figure 6; The second-stage regression equation of Hubei Province is log (R/S)=1.368log(n)-0.814. The fitted curve is shown in Figure 9. It can be found that the H value of both stages is not equal to 0.5, indicating that the newly confirmed number of newly diagnosed coronaviruses in Hubei Province is not random ly walked, which also reflects that a series of tracking and isolation measures adopted by Hubei Province have played a certain role.

The H value in the first stage is 0.574> 0.5, indicating that the sequences are generally long-term memory and are positively correlated, that is, if the number of newly confirmed cases during the previous analysis increases (sequence upwards), then the newly confirmed cases during the next analysis The probability of increase is high. Therefore, it can be predicted from the H value that the number of newly confirmed cases in the short term is still rising. However, from the data point of view, after the number reached 14840 cases on the 12th, the number dropped sharply to 4,823 on the 13th, and the confirmed cases began to show a downward trend. This is due to an inflection point in the trend brought about by changes in diagnostic criteria.

The H value in the second stage is 1.368> 1, which indicates that the sequence is fully predictable. This explains to some extent that under the current environment of national control and concerted national efforts, the epidemic of new crown pneumonia is preventable and controllable.

###### 4.2.2-2 Analysis of Epidemic Situation in Other Provinces

Hubei Province, as an area where the early COVID-19 infections outbroke, has concentrated on studying the transmission mechanism and the trends of confirmed cases to tell the similarities and differences with other provinces in China, and it is of significance to take different protective measures for different provinces. In this article, the reasons for the selection and stage division of the number of newly confirmed cases in other provinces in China except Hubei are the same as above. The daily number of newly confirmed cases from January 23 to February 11 is used as the first stage data. We take the period between February 13 to March 3 as the second stage. Figures 10-15 can be obtained through Matlab data analysis. The results are as follows:

**Figure10.**
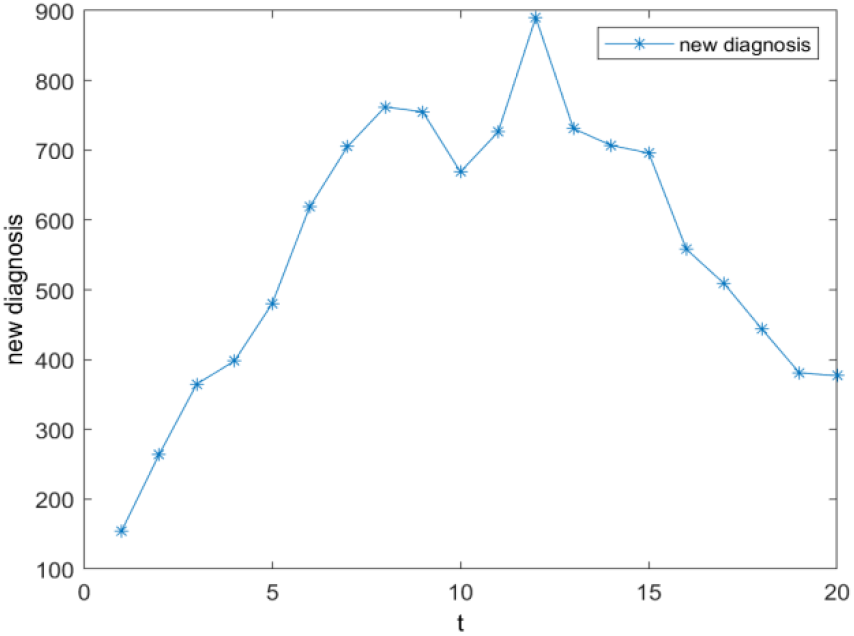
Daily diagnosis in other provinces(first stage)

**Figure11.**
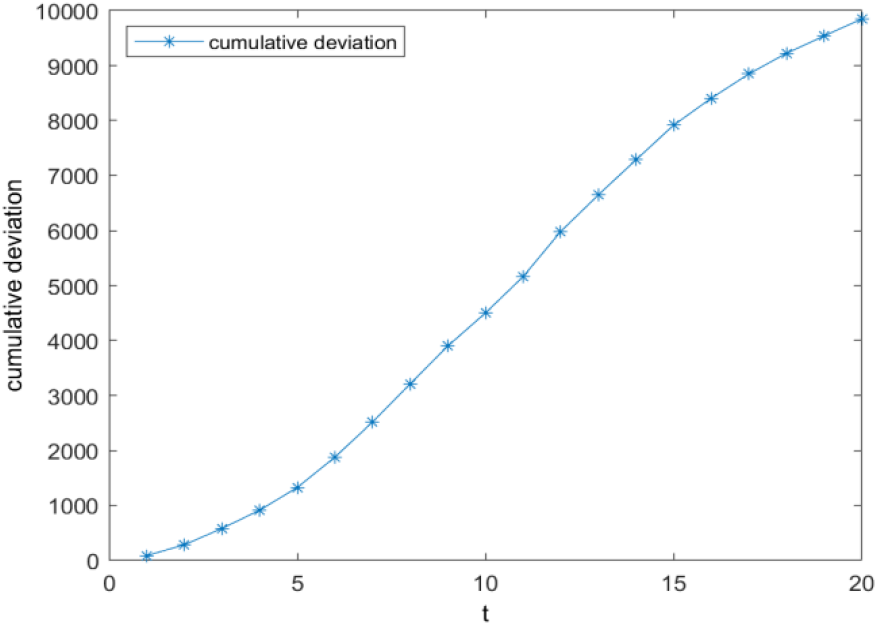
Cumulative deviation in other provinces(first stage)

**Figure12.**
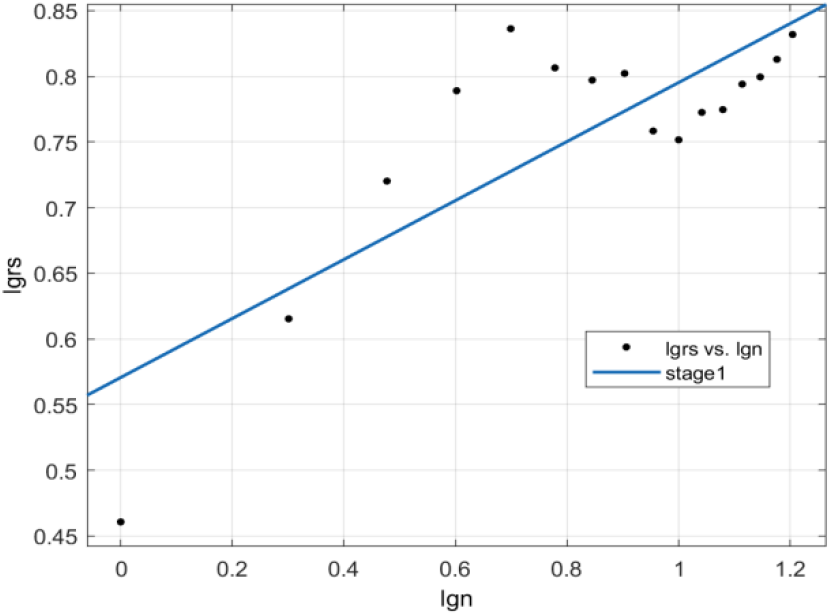
Regression of Log(R/S) to log(n) (first stage)

**Figure13.**
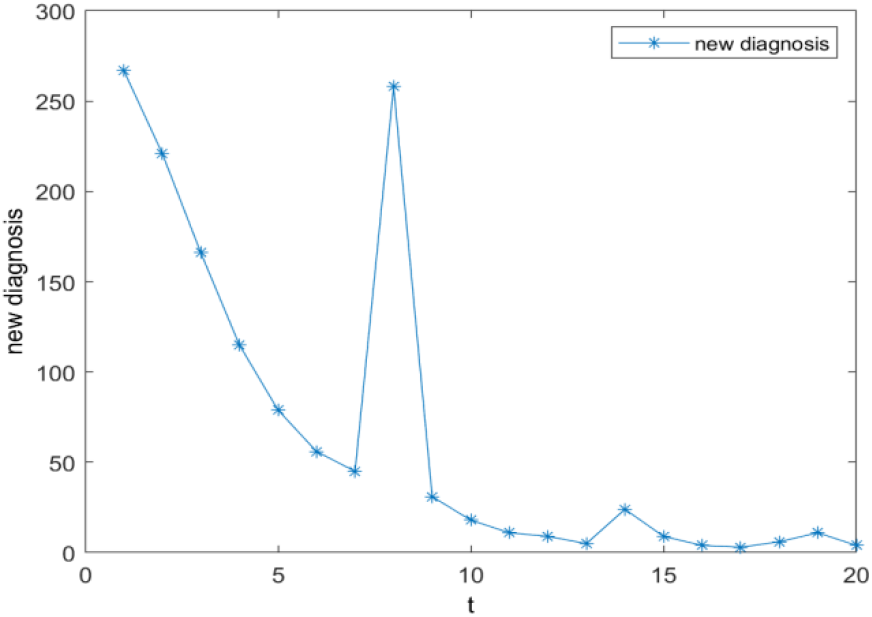
Daily diagnosis in other provinces(second stage)

**Figure 14.**
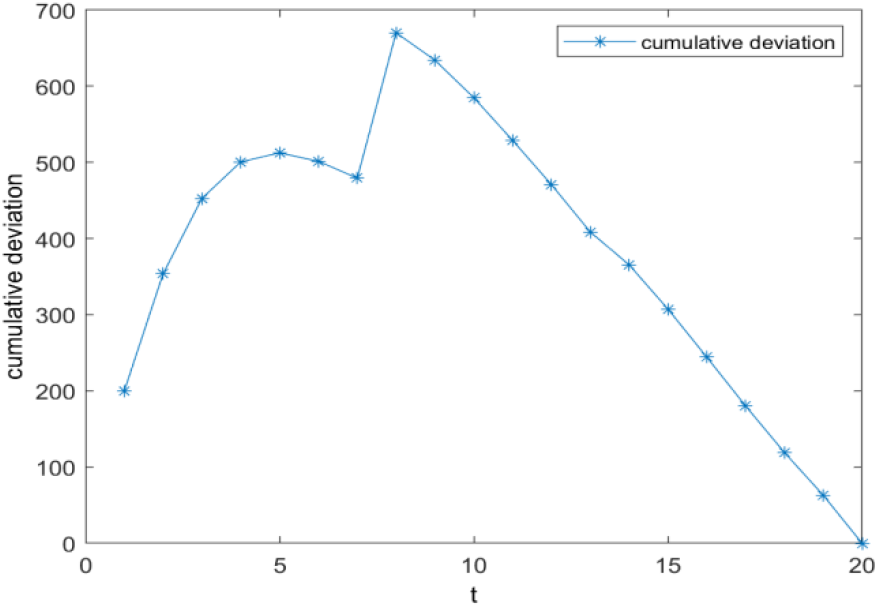
Daily diagnosis in other provinces(second stage)

**Figure15.**
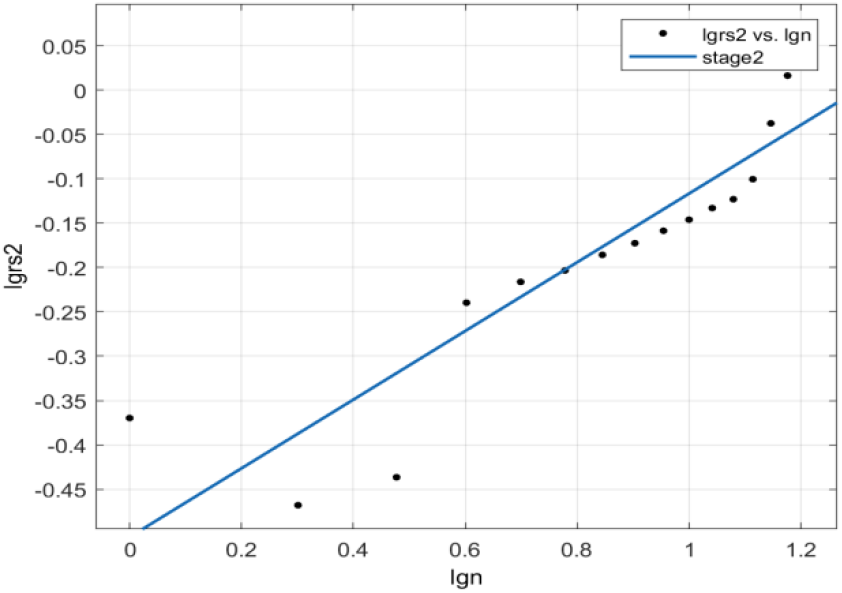
Regression of Log(R/S) to log(n) (second stage)

##### 2.1 Time series characteristics

From the change in the number of the newly confirmed cases in the first stage over time, the curve generally showed a trend of rising before decreasing. Before t = 10, the diagnosis and treatment of COVID-19 infection cases are still in the initial stage. Patients and medical personnel”s understanding of the disease has not reached the level of infectious diseases. The general public has not paid enough attention to it and has not adopted protective measures widely. With the outbreak of the incubation virus, the number of newly diagnosed cases continues to rise, reaching a small peak around February 10. Since February, the number of new cases has increased sharply again, probably because the virus carriers in the population flowing out of Hubei Province from mid-January to January have passed the incubation period and their symptoms have begun to appear. A first-level response to major public health emergencies was initiated. Medical staff and administrative staff efficiently screened out latent and close contacts, and actively adopted preventive and control measures such as isolation, disinfection, and hospitalization. Increasing the efficiency of disease management mechanisms has indirectly led to rise in newly diagnosed cases. Beginning around February 12, local medical systems have basically established a coping mechanism for new crown virus infection. From quarantining patients and tracking close contacts to detection and treatment of patients, methodical prevention and control measures have had a positive effect, and new confirmed cases have been added. The number started to fall.

The time series data of confirmed cases in the second stage fluctuated more obviously than in the previous stage. As shown in Figure 13, the peak occurred on February 13, probably because the national standard for diagnosis has changed from nucleic acid testing to clinical diagnosis from February 11. Many previously undiagnosed patients were included in new cases. This growth peak lasted 2-3 days, and returned to its previous path after February 15. Except for the peak point, the daily confirmed new cases in the second stage showed a downward trend, and it can be seen that the early anti-epidemic effect has gradually emerged. After February 25, the number of newly confirmed cases per day remained basically in the single digits.

##### 2.2 Cumulative Deviation

It can be seen from Figure 11 that the cumulative deviation curve of the first stage in other provinces across the country slopes to the upper right, and the slope increases first and then decreases, indicating that the daily new confirmed cases in this stage are generally larger than the serial mean. During the period from January 23 to February 11, although the epidemic prevention and control has achieved initial results, its development still shows an expanding trend and has not yet entered a period of decline. It can be seen from FIG. 14 that the cumulative deviation curve in the second stage shows an upward trend and then a downward trend, and reaches a peak around February 21. The daily new cases before the peak point are generally larger than the overall mean value, and then start to be below the mean value point, reflecting that the newly confirmed cases have been initially contained.

#### 3. 2.3 Fractal Hurst Index

We obtained the regression equation log (R / S) = 0.2246log (n) +0.571 using Matlab to fit the first stage data. It can be seen that H = 0.2246, and the fitting curve is shown in Figure 12, which shows that the scattered points follow the fractal Brownian motion process. Generally, the random walk sequence spreads more slowly, and the sequence points tend to return to historical points in the future. The fitting regression equation in the second stage is log (R / S) = 0.3872log (n) -0.504, H = 0.3872. The fitting curve is plotted in Figure 15, which presents that the daily new cases in this stage show a downward trend. We do not fully understand the mechanism of virus development and mutation, yet we cannot relax our vigilance. With the development of foreign epidemics, new imported cases may appear in China, bringing new growth peaks. Strict prevention and screening measures are needed for this.

In general, the H values of the two stages in other provinces are less than 0.5, and the time-series data of newly confirmed diagnosis nationwide may return to historical points in the future. The number of newly added people in the first stage has a peak on the 9th and 12th days, respectively. The growth rate of newly diagnosed patients has slowed in the last five days.

According to the prediction of the H index, the number of new people will pick up in a period of time after the first stage. It then gradually declined and the number of new diagnoses will gradually decrease after the second stage. The data shows that 377 people were newly diagnosed on February 11, and the number of people increased sharply on February 12 due to changes in clinical diagnostic standards, while 267 people were added on February 13, and the overall decline was subsequently shown. This fits the second stage H index.The time series data showed a high peak on February 20, which may be related to the occurrence of clustered morbidity events in some areas, such as the confirmed diagnosis of prison groups.

## 5 Conclusion

This paper establishes a rescaled range analysis model to analyze and predict the epidemic situation in three different areas. The obtained H index is located in different intervals. Using this result we can see that there are also differences in the development paths among regions with different outbreak times both at home and abroad. By analyzing historical data, we can make a trend prediction of the future situation of virus transmission.

The results of the model analysis show that the Hurst value of the new change rate in Italy is greater than 1, and the time series follow a fractal Brownian motion, indicating that the future trend is similar to the past with a high probability. Therefore, the Italian government should focus on controlling the infection in terms of source, cutting off the transmission route, following the principle of early detection, early isolation, and early treatment to reduce the gathering of large people, in the meantime regularly cleaning of public spaces is necessary. The number of newly diagnosed patients in Italy is highly similar to that in Hubei. It can learned from the strict measures adopted by Hubei in isolation and treatment, community prevention and other measures. Also, they can adopt effective protection measures based on the actual situation of the epidemic in the country to significantly reduce the population contact rate. Blocking the route of transmission, and thoroughly controlling the spread of the epidemic can also help. The H indexes of all provinces in China except Hubei are in the range of (0,0.5), indicating that the virus transmission has fractal Brownian motion characteristics. There may be a correlation between the future path and historical data, and the number of newly diagnosed patients will be increased in the short term. We can see the risk of returning to the historical path. Therefore, the masses and medical staff need to take corresponding preventive and controlling measures. Yet it is not time to relax our vigilance, we should continue to strengthen our protection and observe the changes in data, as well as pay particular attention to the cyclical fluctuations that rise after the number of confirmed diagnoses declines.

## Data Availability

National Health Commission of the People's Republic of
China and WHO

